# Changes in Alcohol Consumption during the COVID-19 Pandemic: Evidence from Wisconsin

**DOI:** 10.1101/2022.11.07.22282029

**Authors:** Rachel Pomazal, Laura McCulley, Amy Schultz, Noah Stafford, Mikayla Schowalter, Kristen Malecki

## Abstract

**Introduction:** The COVID-19 pandemic increased stress levels broadly in the general population. Patterns of alcohol consumption are known to increase in times of increased stress like natural disasters, disease outbreaks, and economic turmoil. Wisconsin is an important place to study changes in alcohol consumption because it is one of the heaviest-drinking states in the United States. The primary aim of this study is to identify changes in alcohol use at three distinct timepoints during the COVID-19 pandemic in a statewide sample.

**Methods:** An online survey was sent to 5,502 previous Survey of the Health of Wisconsin (SHOW) participants to ask about a wide range of topics related to COVID-19. The timepoints were taken May through June 2020 (Wave 1), January to February 2021 (Wave 2), and June 2021 (Wave 3) The sample included 1,290, 1,868, and 1,585 participants in each of the three waves respectively. Changes in alcohol consumption (whether they drank more, about the same, or less) were examined by race, age, gender, educational attainment, annual income, anxiety and depression status, remote work status, whether the participant experienced employment changes due to COVID-19, and whether there were children present in the home. Within-wave univariate changes in alcohol consumption were evaluated by demographics using a chi-squared test.

**Results:** In all three waves, those with anxiety, a bachelor’s degree or higher, two younger age groups, and those with children in the home were significantly more likely to increase alcohol consumption. Those reporting depression, those in the highest income quartile, and those working remotely were more likely to report increased drinking in the first two surveys, but not in the third survey. Participants reporting changes in employment due to COVID-19 were more likely to increase drinking in the first survey only. Non-white participants were more likely to report decreased drinking in the first survey only.

**Conclusions:** There may be subpopulations in Wisconsin at higher risk for the negative effects of heavy drinking during the pandemic like those with anxiety, those with children in the home, those with a bachelor’s degree or higher, and those in younger age groups, as these groups had consistently higher alcohol use that did not subside 15 months after lockdowns began.

## Introduction

Early in the COVID-19 pandemic social restrictions effectively reduced viral transmission; however, they also introduced a host of new risks including changes in personal behaviors including alcohol consumption. Increased stress and social isolation (Holmes et al., 2020), employment and other economic changes were highly prevalent, and continued for the next 24 months (Kornreich, 2022). Research conducted prior to COVID-19 has connected psychological distress to problematic alcohol consumption (Bott et al., 2005; Markman Geisner et al., 2004; Okoro et al., 2004; Brière et al., 2014; Bolton et al., 2006; Robinson et al., 2009). Further, social isolation (Fairbairn and Sayette, 2014) and stress (Keyes et al., 2012; Clay and Parker, 2020) are important psychological factors predicting disordered drinking. Increased stress and anxiety tend to increase substance use as a coping mechanism (Baker et al.; 2004; Goldmann and Galea, 2014); which is exacerbated during natural disasters, pandemics, and similar high-stress or traumatic experiences (Cepeda et al., 2010; Bor et al., 2013; Wu et al., 2008; Boscarino et al., 2011; Richman et al., 2008; Kanehara et al., 2016). Thus, an increased understanding of how alcohol patterns and behaviors changed across the first twelve months of the COVID-19 pandemic would offer some important insights.

Patterns of alcohol consumption are culturally specific and little information was available early in the pandemic. A survey of adults living in Hong Kong during the 2003 SARS-CoV outbreak found that 6.8% of randomly sampled adults, and 6% of hospital employees reported increased alcohol use as a coping strategy (Lau et al., 2005; Wu et al., 2008). An early study conducted via survey during the SARS-CoV-2 pandemic lockdowns in China found increased levels of depression and anxiety in a snowball sample of the general public via university students (Yao et al., 2020). Other studies showed a greater increase in alcohol consumption as a coping strategy during the initial lockdown phase of the pandemic (Lee et al., 2020; Lechner et al., 2020; Pollard et al., 2020), which is consistent with previous findings on the psychological impact of pandemic-related quarantines (Brooks et al., 2020). In the U.S. alcohol sales in early March to mid-April 2020 rose significantly with an increase in liquor store sales of 54% and online alcohol sales of 262%, compared to 2019 data (Bremner, 2020). Little data is available on changing trends in alcohol consumption at a state level, in places like Wisconsin, an upper Mid-western state of the United States.

Currently in the U.S. nearly one in six adults engages in heavy episodic drinking (HED) four times per month, where they consume, on average, seven drinks per episode (SAMHSA, 2018, Kanny et al., 2018). Aside from acute consequences of heavy alcohol use, such as alcohol poisoning and blackouts (White et al., 2018), there is the potential that, longer-term alcohol misuse may escalate into a clinical alcohol use disorder (Griswold et al., 2018) or comorbidities between mental health and substance use disorders (Farrell et al., 2003; Jane-Llopis and Matytsina, 2006; Lai et al., 2015).

Consequences of alcohol misuse can be severe. In the U.S, alcohol misuse is the fourth leading preventable cause of death (Starhe, 2014),. Excessive drinking is also a risk factor for numerous chronic conditions including high blood pressure, heart disease, and stroke (Centers for Disease Control and Prevention, 2022) and is associated with increased violence (Snowden, 2019), crime (Toomey et al., 2012), poverty (Khan et al., 2002), sexually transmitted diseases (Cook & Clark, 2005), and other public health harms.

In Wisconsin alcohol overconsumption has been a persistent public health burden. Wisconsin has been consistently identified as one of the heaviest-drinking states in the US, and has an adult population that is more likely to drink alcohol (64%) than the national average (55%) (Dwyer-Lingren et al., 2015; Wisconsin Department of Health Services, 2022). Wisconsin ranks third in the nation for adult binge drinking, defined as consuming more than 4 drinks for women or 5 drinks for men on an occasion, (22% of Wisconsinites compared to 16% national average) (Centers for Disease Control and Prevention, 2017). Wisconsinites were less likely (38%) than the national average (45%) to perceive significant risk from weekly binge drinking (Centers for Disease Control and Prevention, 2017; Wisconsin Department of Health Services, 2022). Given this baseline of high drinking, and low perception of alcohol consumption risks as part of the culture, public health officials in Wisconsin were weary of an additional spike in alcohol use in response to stress incurred due to the COVID-19 pandemic.

### Study Aims

The primary aim of this study is to identify changes in alcohol use at three distinct timepoints during the COVID-19 pandemic in a statewide sample. This study also aimed to identify and understand potential public health implications of any changes in alcohol use.

## Methods

### The Survey of the Health of Wisconsin (SHOW)

The Survey of the Health of Wisconsin (SHOW) is a statewide population-based health study which began in 2008 to inform health research in Wisconsin through data collection on a range of health, economic, and environmental questions in a representative sample of the state, including objective body measurements and biosamples, as well as self-reported data from computer assisted personal interviews and self-administered questionnaires (Nieto, et al. 2010). SHOW currently includes 5,846 adult participants recruited between 2008 and 2019, including an intentional oversampling from historically underrepresented populations in biomedical research including African Americans and Hispanics in the City of Milwaukee, Wisconsin. More details about the SHOW cohort, sampling frame and study design are available elsewhere (Malecki et al., 2022a).

### The SHOW COVID-19 Community Impact Survey

#### Study Participants and Recruitment

In spring of 2020, The SHOW developed the online COVID-19 Community Impact Survey in collaboration with over 25 professors and investigators across the University of Wisconsin, Madison. The survey was administered at three different timepoints over the course of 2020-2021 (referred to as “waves” of the survey). The survey aimed to capture COVID-19 perceptions, beliefs and behaviors, as well as how the pandemic affected their mental, physical and emotional health and their life overall. The online survey was administered May through June 2020 (Wave 1), January to February 2021 (Wave 2), and June 2021 (Wave 3) (“Covid-19 Public Use Data”, 2022). SHOW participants were eligible to participate in any or all three waves if they had consented to be contacted for future research and have provided an email or phone number. Among the 5,846 adult SHOW cohort, n=5,502 met eligibility criteria and were invited to participate in every wave of survey.

A unique web-based survey link was emailed to all eligible participants with information on what the survey would ask. The survey was administered online via UW ICTR-CAP REDcap. Participants were also contacted by phone if they did not have a valid email address and were asked for a valid email address at that time, or had the opportunity to complete a shortened version of the survey via phone interview.

The study was approved by the University of Wisconsin-Madison Health Science Institutional Review Board. All participants who completed the online COVID-19 or telephone surveys received a $25 electronic gift card.

In total, 1,403 participants completed the Wave 1 survey, 1,889 participants completed the Wave 2 survey, and 1,615 participants completed the 1-year follow up Wave 3 survey (Malecki et al, 2021). Information on how many participants completed each survey, and how many participants completed multiple surveys are available on the SHOW website (“Covid-19 Public Use Data”, 2022). Additionally, n=55 completed the telephone survey. More details about the SHOW COVID-19 Community Impact Survey and the cohort and methods have been described elsewhere (Malecki et al., 2022b) and are available on the SHOW website.

For this study, only participants who completed the online survey were included in analyses; those who completed the telephone interview survey were excluded. A total of n=1,290, n=1,868, and n=1,585 had complete data on alcohol consumption, and were included in analysis for waves 1, 2, and 3, respectively.

### Demographics

Gender, income, educational attainment, presence of children in the home, smoking status, remote work status, changes in employment during the COVID-19 pandemic, anxiety and depression status were self-reported within the survey. Anxiety and depression status were determined by asking if participants had ever been told by a doctor or health care professional that they had these conditions that were not related to COVID-19. Self-reported race was collected in four categories, then was categorized as non-Hispanic white and non-white due to a relatively small number of non-white participants in the COVID-19 Impact surveys. Age at time of survey was analyzed categorically as 21-39, 40-59, 60-74, and 75 years of age and older to group participants into relevant generational cohorts that may differ in drinking habits. Income groups were determined by self-reported annual household income less than $29,999, between $30,000-$59,999, between $60,000-$99,999 and greater than $100,000. Health status was assessed on a 5-point Likert scale based on the validated SF-12 health survey with possible responses Excellent, Very Good, Good, Fair, or Poor. These were then grouped into 3 categories: Fair/Poor, Good, and Excellent/Very Good for ease of analysis. Educational attainment was grouped by High School/G.E.D or less, Some College, and Bachelor’s Degree or Higher to ascertain relevant cut points in average earning potential.

### Statistical Analysis

All statistical analysis was completed in SAS v9.4. Participants were asked to self-report whether their alcohol consumption was “a lot more, a little more, about the same, a little lower, or much lower” in the last 60 days compared to before the pandemic in the first wave, and in subsequent waves, the question was asked cross-sectionally compared to before the pandemic (since July 1, 2020 or since February 1, 2021). We then categorized these responses into whether participants drank more, about the same, or less than before the pandemic to ensure sufficiently large sample size in each category. Only those who completed questions related to alcohol consumption were included in analysis. Those with incomplete demographic data were included in the comparisons they completed, and were not entirely excluded. Everyone that completed a wave was included in analyses for that time point, regardless of participation in other waves. Within-survey univariate differences in changes in alcohol consumption were compared using a chi-squared test. Differences in alcohol consumption were evaluated by gender, age group, race, income, anxiety and depression status, health status, remote work status, whether the participant experienced changes in employment, and presence of children in the home as these were found to be significant in other literature.

## Results

In the first survey n=1,290 had complete data on alcohol consumption, in the second survey, n=1,868 had complete data on alcohol consumption, and in the third survey n=1,585 had complete data on alcohol consumption for this analysis Table 1 describes the demographics of the sample, including differences in changes in alcohol consumption. All timepoints were majority non-Hispanic white, female, with a bachelor’s degree or higher. In Wave 1, 23.18% of respondents reported increased drinking; in Wave 2, 18.84% of respondents reported increased drinking, and 15.33% of respondents reported increased drinking in Wave 3.

|  | **Wave 1**<br>**(n=1,290)** | **Wave 2**<br>**(n=1,868)** | **Wave 3**<br>**(n=1,585)** |
| --- | --- | --- | --- |
| **Gender** |  |  |  |
| Male | 464 | 725 | 593 |
| Female | 817 | 1129 | 978 |
| **Age** |  |  |  |
| 21-35 yrs | 151 | 175 | 139 |
| 36-55 yrs | 422 | 608 | 484 |
| 56-75 yrs | 614 | 883 | 787 |
| Greater than 75 yrs | 101 | 201 | 163 |
| **Race** |  |  |  |
| White (Non-Hispanic) | 1139 | 1624 | 1371 |
| Non-White | 149 | 242 | 189 |
| **Education** |  |  |  |
| H.S./GED or Less | 197 | 301 | 246 |
| Some College | 411 | 648 | 555 |
| Bachelors or Higher | 675 | 912 | 769 |
| **Income** |  |  |  |
| < \$30,000 | 163 | 252 | 208 |
| \$30,000 - \$59,999 | 301 | 447 | 362 |
| \$60,000 - \$99,999 | 347 | 498 | 417 |
| > \$100,000 | 364 | 480 | 431 |
| **Self Reported Health** |  |  |  |
| Excellent or Very Good | 783 | 1091 | 939 |
| Good | 393 | 609 | 484 |
| Fair or Poor | 113 | 167 | 160 |
| **Children in Home** |  |  |  |
| Children Present | 379 | 528 | 401 |
| No Children Present | 911 | 1340 | 1184 |

In all three survey timepoints, changes in alcohol consumption varied significantly with anxiety status where 30.7%, 24.9%, and 20.3% of those with anxiety increased drinking in each wave, compared to 21.4%, 17.3%, and 14.1% respectively (p=0.0066, 0.0027, and 0.0248). 27.7%, 21.5%, and 18.9% of those with a bachelor’s degree or higher increased drinking in each wave, compared to those with a high school education or less (15.2%, 15.0%, or 13.4% respectively), or those with some college education (20.0%, 16.8%, or 11.2% respectively) (p=0.0016, <0.0001, and 0.0003). Those in the younger two age categories were nearly twice as likely to increase drinking in all waves compared to those aged 60-74 years, and at least three times as likely to increase drinking compared to those aged 75 years and up (p<0.0001 in the first and third surveys, and p=0.0015 in the second survey). Participants with children in the home were more likely to increase drinking habits all three surveys (34.6%, 25.6%, and 21% versus 18.4%, 16.2%, and 13.4% respectively) (p<0.0001 in the first two surveys, and p=0.0013 in the third survey). Those reporting depression (p=0.0224, 0.0261, and 0.3), working remotely (p<0.0001 and 0.0702), and those in the highest income quartile (p=0.0001 and 0.09) were significantly more likely to report increased drinking habits in the first two surveys, but not the third survey. Participants reporting changes in employment were significantly more likely to report increased drinking at the first timepoint, but not the second or third timepoints (p=0.0387, 0.87, and 0.99). White participants were more likely to report similar drinking behaviors at the first timepoint, and non-white participants were more likely to report decreased drinking behaviors at the first timepoint, but results were similar at subsequent timepoints (p=0.0006, 0.78, and 0.30). Those reporting Fair/Poor health at the second timepoint were less likely to report increased drinking than those reporting better health statuses (p=0.0233). See Table 2 for a complete list of within-survey comparisons.

**Table 2.** Changes in Alcohol Consumption by Demographics

|  | Wave I |  |  |  | Wave II |  |  |  | Wave III |  |  |  |
| --- | --- | --- | --- | --- | --- | --- | --- | --- | --- | --- | --- | --- |
|  | Drank<br>More (%) | Drank the<br>Same (%) | Drank<br>Less (%) | p-trend | Drank<br>More (%) | Drank the<br>Same (%) | Drank<br>Less (%) | p-trend | Drank<br>More (%) | Drank the<br>Same (%) | Drank<br>Less (%) | p-trend |
| **Race** |  |  |  |  |  |  |  |  |  |  |  |  |
| White | 23.09 | 62.86 | 14.05 | 0.0006 | 18.72 | 64.35 | 16.93 | 0.7832 | 15.83 | 70.09 | 14.08 | 0.2995 |
| Non-White | 24.16 | 50.34 | 25.5 |  | 19.83 | 64.88 | 15.29 |  | 11.64 | 72.49 | 15.87 |  |
| **Gender** |  |  |  |  |  |  |  |  |  |  |  |  |
| Male | 20.91 | 64.66 | 14.44 | 0.1671 | 17.93 | 63.72 | 18.34 | 0.2587 | 13.66 | 72.01 | 14.33 | 0.4292 |
| Female | 24.6 | 59.36 | 16.03 |  | 19.57 | 64.92 | 15.5 |  | 16.05 | 69.63 | 14.31 |  |
| **Income** |  |  |  |  |  |  |  |  |  |  |  |  |
| <\$29,999 | 17.79 | 58.28 | 23.93 | 0.0001 | 15.08 | 66.27 | 18.65 | 0.0001 | 12.5 | 72.6 | 14.9 | 0.0913 |
| \$30,000-\$59,999 | 18.94 | 65.12 | 15.95 | | 18.12 | 66.44 | 15.44 | | 13.81 | 70.99 | 15.19 | |
| \$60,000-\$99,999 | 23.92 | 64.84 | 11.24 | | 17.87 | 67.87 | 14.26 | | 16.55 | 70.74 | 12.71 | |
| >\$100,000 | 30.22 | 54.95 | 14.84 | | 25.42 | 54.17 | 20.42 | | 19.72 | 63.81 | 16.47 | |
| **Anxiety Status** |  |  |  |  |  |  |  |  |  |  |  |  |
| Anxiety | 30.68 | 54.58 | 14.74 | 0.0066 | 24.93 | 60.98 | 14.09 | 0.0027 | 20.33 | 66.89 | 12.79 | 0.0248 |
| No Anxiety | 21.37 | 63.14 | 15.5 |  | 17.34 | 65.24 | 17.41 |  | 14.14 | 71.25 | 14.61 |  |
| **Depression Status** |  |  |  |  |  |  |  |  |  |  |  |  |
| Depression | 30.13 | 56.33 | 13.54 | 0.0224 | 23.77 | 60.93 | 15.3 | 0.0261 | 17.35 | 70.75 | 11.9 | 0.3065 |
| No Depression | 21.68 | 62.58 | 15.74 |  | 17.64 | 65.25 | 17.11 |  | 14.87 | 70.33 | 14.79 |  |
| **Remote Work Status** |  |  |  |  |  |  |  |  |  |  |  |  |
| Remote Work | 35.17 | 52.91 | 11.93 | <.0001 | 26.41 | 54.52 | 19.07 | <.0001 | 20.9 | 61.94 | 17.16 | 0.0702 |
| No Remote Work | 19.11 | 64.38 | 16.51 |  | 16.72 | 67.17 | 16.11 |  | 14.82 | 71.19 | 13.99 |  |
| **Health Status** |  |  |  |  |  |  |  |  |  |  |  |  |
| Excellent/Very Good | 23.37 | 62.45 | 14.18 | 0.3322 | 20.16 | 62.97 | 16.87 | 0.0233 | 16.72 | 68.8 | 14.48 | 0.0968 |
| Good | 24.43 | 59.03 | 16.54 |  | 19.05 | 65.19 | 15.76 |  | 13.64 | 70.87 | 15.5 |  |
| Fair/Poor | 17.7 | 62.83 | 19.47 |  | 9.58 | 70.66 | 19.76 |  | 12.5 | 78.13 | 9.38 |  |
| **Education** |  |  |  |  |  |  |  |  |  |  |  |  |
| HS/GED or less | 15.23 | 67.51 | 17.26 | 0.0016 | 14.95 | 70.43 | 14.62 | <.0001 | 13.41 | 74.39 | 12.2 | 0.0003 |
| Some College | 19.95 | 63.26 | 16.79 |  | 16.82 | 68.36 | 14.81 |  | 11.17 | 75.68 | 13.15 |  |
| Bachelor's Degree or Higher | 27.7 | 58.37 | 13.93 |  | 21.49 | 59.87 | 18.64 |  | 18.86 | 65.28 | 15.86 |  |
| Age Group |  |  |  |  |  |  |  |  |  |  |  |  |
| 21-39 | 33.11 | 47.02 | 19.87 | <.0001 | 26.86 | 57.14 | 16 | 0.0015 | 18.71 | 63.31 | 17.99 | <.0001 |
| 40-59 | 34.36 | 52.37 | 13.27 |  | 26.32 | 57.57 | 16.12 |  | 22.31 | 64.26 | 13.43 |  |
| 60-74 | 16.45 | 68.89 | 14.66 |  | 14.95 | 68.29 | 16.76 |  | 12.2 | 73.44 | 14.36 |  |
| >75 | 2.97 | 76.24 | 20.79 |  | 6.47 | 74.13 | 19.4 |  | 6.75 | 79.14 | 14.11 |  |
| Employment Change During COVID-19 |  |  |  |  |  |  |  |  |  |  |  |  |
| Changes in Employment | 25.36 | 59.52 | 15.12 | 0.0387 | 19 | 64.03 | 16.97 | 0.8673 | 15.26 | 70.55 | 14.18 | 0.9914 |
| No Changes in Employment | 19.11 | 65.11 | 15.78 |  | 18.45 | 65.31 | 16.24 |  | 15.41 | 70.25 | 14.34 |  |
| Presence of Children in the Home |  |  |  |  |  |  |  |  |  |  |  |  |
| Children in Home | 34.56 | 51.98 | 13.46 | <.0001 | 25.57 | 59.66 | 14.77 | <.0001 | 20.95 | 66.58 | 12.47 | 0.0013 |
| No Children in Home | 18.44 | 65.42 | 16.14 |  | 16.19 | 66.27 | 17.54 |  | 13.43 | 71.71 | 14.86 |  |

## Discussion

To our knowledge this is the first study to examine changes in alcohol consumption during several phases of the COVID-19 pandemic in Wisconsin. By utilizing serial surveys, we are able to examine changes in these dynamics within a relatively short period of time. Wisconsin is an opportune state to study these changes during the pandemic because of its strong culture of drinking. According to the Behavioral Risk Factor Surveillance System Wisconsin ranks third in adult binge drinking (Centers for Disease Control and Prevention, 2022). Therefore, Wisconsinites may be at higher risk for increased drinking behavior in stressful situations, like a global pandemic. Statewide surveys allow us to get a clearer picture of the impact of COVID-19 on regions and communities across the state during a time when in-person data collection was difficult.

We examined associations based on previously published literature on changes in alcohol consumption during the COVID-19 pandemic including gender, race, reported depression or anxiety, remote work, presence of children in the home, and others. We found increased drinking habits among those reporting anxiety at all three timepoints, and among those reporting depression during the first two timepoints. During the pandemic, these participants may have had lower access to therapies and other interventions to treat anxiety, and may have self-medicated with alcohol instead. Several other studies have examined changes in alcohol consumption and other substance use during the COVID-19 pandemic in various contexts. Several studies found significant associations between increased alcohol consumption and those reporting anxiety and depression symptoms (Lechner et al., 2020; Knell et al., 2020; Avery et al., 2020). At all three timepoints, we also found increased drinking behavior in those reporting children in the home. There is no doubt that the presence of children during the pandemic, especially with remote schooling, may have increased stress levels of adults in the household. Grossman et al also reported increased drinking habits in those reporting children in the home among adults in the US. We also found increased drinking at all three timepoints among younger age groups and those with a bachelor’s degree or higher. This may be because younger people are more involved in the drinking culture of Wisconsin. Those with a bachelor’s degree or higher may have higher socioeconomic status on average and work from home, or may feel that increased drinking behavior is more acceptable than those with lower educational attainment. They may also be more able to access alcohol due to increased means to purchase alcohol when there are pandemic-related financial strains, and other measures of access to alcohol. Rolland et al similarly found increases in alcohol use among younger age groups, higher educational attainment, and current psychiatric treatment among the general population of France, which mirrors our results. Conversely, several studies found women were more likely to increase drinking habits (Dumas et al., 2020; Rodriguez et al., 2020). Both studies utilized different metrics to assess alcohol consumption from those used here (number of alcohol using days, binge drinking, and number of drinks per drinking occasion versus self-reported changes in drinking habits), which may account for some of these differences. Additionally, these studies were conducted at only one timepoint, so it is unclear whether these results would hold had the survey been completed multiple times at different phases of the pandemic. Finally, The study by Dumas et al was conducted in Canadian adolescents, who may have different drinking habit changes compared to adults in the US due to differing attitudes toward adolescent drinking in both countries, and due to general age group differences.

This study has several strengths and limitations that may impact the results of the survey. First, 986 participants completed the survey at all three timepoints, and 1,675 participants completed at least two timepoints. This repeated participation enables us to examine changes in alcohol consumption through different phases of the pandemic in the same participants. Additionally, the rich survey data collected allows us to explore many important associations with changes in alcohol consumption. Since Wisconsin is such an advantageous place to study alcohol consumption, it is a particular strength of this study to have conducted this work here. A limitation of this study is the need to combine all non-white race and ethnicity groups into one, as there were insufficient responses within each race and ethnicity group to draw reliable conclusions. The wording of some questions changed slightly between timepoints of the survey, which may have impacted participants’ responses. These surveys also relied on self-reports of demographics, as well as changes in behavior over time, which is vulnerable to recall bias. Clear, objective definitions of increased or decreased drinking behaviors were not defined within the survey, which relies on participants’ interpretations of the question. Additionally, when asking about potentially sensitive topics like changes in alcohol consumption, employment, income, health status, and diagnoses of anxiety and depression, social desirability bias may be important. Participants may under-report these factors, which may have resulted in differential misclassification to ‘healthier’ statuses. Finally, since the survey was conducted online, past SHOW participants who do not have internet access or could not complete the online survey for other reasons could not be included in the data. This may skew the data as internet access may be related to certain demographics and may be related to changes in alcohol consumption throughout the pandemic.

Our study suggests that certain groups may have been differentially at risk for increased alcohol consumption during various phases of the COVID-19 pandemic, which may put them at elevated risk for adverse health outcomes. More research is needed to understand the scope of alcohol and substance use changes among Wisconsinites occurring in the constantly changing phases of the pandemic. Healthcare providers should pay special attention to patients with anxiety, younger patients, and patients with children in the home and consider discussing risks of increased alcohol consumption with them. Future studies should examine differences in alcohol consumption changes between pandemic phases, going beyond the within-phase comparisons here. These studies should also use rich survey data provided by SHOW to link COVID-19 Impact data with other important exposures like housing, geography, residential history, and biological samples to better understand these dynamics in Wisconsin. Finally, longitudinal follow-up on the impacts of COVID-19 among these participants should be conducted as Wisconsinites change the ways in which they interact with the virus long-term.

## Data Availability

All data produced in the present study are available upon reasonable request to the authors. A public use version of this dataset is available online at https://show.wisc.edu/data/covid-19-public-use-data/

